# Attitudes and Perceptions of Medical Researchers Towards the Use of Artificial Intelligence Chatbots in the Scientific Process: A Large-Scale, International Cross-Sectional Survey

**DOI:** 10.1101/2024.02.27.24303462

**Authors:** Jeremy Y. Ng, Sharleen G. Maduranayagam, Nirekah Suthakar, Amy Li, Cynthia Lokker, Alfonso Iorio, R. Brian Haynes, David Moher

## Abstract

**Background:** Chatbots are artificial intelligence (AI) programs designed to simulate conversations with human users through text or speech. The use of artificial intelligence chatbots (AICs) in scientific research presents benefits and challenges. Although the stances of journals and publishing organizations on AIC use is increasingly clear, little is known about researchers’ perceptions of AICs in research. This survey study explores attitudes, familiarity, perceived benefits, limitations, and factors influencing adoption of AIC by researchers.

**Methods:** A cross-sectional online survey of published researchers was conducted. Corresponding authors and their e-mail addresses were identified by querying PubMed for articles (any type) published in a MEDLINE indexed journal in the most recent two months and using R script on PubMed metadata. e-Mail invitations were sent to 61560 study authors. The survey, administered on SurveyMonkey, opened on July 9, 2023, and closed on August 11, 2023. Respondents had 3 weeks to complete the survey and were sent 2 reminder e-mails during the weeks of July 17, 2023, and July 24, 2023.

**Results:** 2165 respondents completed the survey (4.0% response rate; 94% completion rate of those who responded). Most were familiar with the concept of AICs (n=1294/2138, 60.5%). About half had used an AIC previously for purposes relating to the scientific process (n=1107/2125, 52.1%). Only 244/2137 (11.4%) respondents reported that their institution offered training on using AI tools of whom 64/244 (26.2%) completed the training. 211/2131 (9.9%) reported that their institution implemented policies regarding AIC use in the scientific process. Most respondents expressed interest in learning more and receiving training on AIC use in the scientific process (n=1428/2048, 69.7%). Respondents had mixed opinions about the potential benefits of using AICs, whereas most agreed on their cons/challenges. Respondents agreed AICs were most beneficial in reducing the workload and administrative burden on researchers (n=1299/1941, 66.9%) and they were most concerned about the lack of understanding behind how AICs make decisions and generate responses (n=1484/1923, 77.2%).

**Conclusions:** Most respondents are familiar with AICs and half used AICs in their own research. Although there is clear interest in understanding how AICs can be used, many hesitate due to existing limitations. Little formal instruction on using AICs is available across academic institutions.

## Background

Artificial intelligence (AI) broadly refers to the ability of a computer or a computer-controlled robot to perform tasks typically associated with human intelligence and its cognitive functions (Copeland, 2023; Petersson et al., 2022; Schroer, n.d.). In medicine and healthcare, AI systems have reduced diagnostic and therapeutic errors, promoted and increased physical activity, and are estimated to have reduced healthcare costs (Ali et al., 2023; Nadarzynski et al., 2019; Palanica et al., 2019). AI has also become an increasingly important tool in scientific research where large amounts of data need to be retrieved, analysed, and interpreted.

Chatbots are AI programs designed to simulate conversation with human users through text or speech (Adamopoulou & Moussiades, 2020; Palanica et al., 2019). Popular AICs include ChatGPT, Bing Chat, YouChat, and Google Bard (Google, n.d.; Microsoft, n.d.; OpenAI, n.d.; You.com, n.d.). In scientific research, AICs can automate tasks such as literature searches and reviews, data analyses and interpretations of large datasets, experimental design, and manuscript writing (Nature, 2023; Sallam, 2023; Salvagno et al., 2023). They also improve the readability of scientific articles for non-native speakers, potentially increasing equity and addressing barriers to promote research diversity (Liebrenz et al., 2023; Sallam, 2023; Salvagno et al., 2023). AI systems can also be trained to differentiate between reproducible and non-reproducible studies by estimating a study’s likelihood of replication (Yang et al., 2020). Overall, AICs improve the efficiency, accuracy and reproducibility crisis in scientific research (Yang et al., 2020).

However, the use of AICs in scientific research has challenges and limitations that need to be addressed. Some major concerns include the accuracy and reliability of AICs in performing or reporting scientific tasks, and ethical issues regarding the use of AI in research. Ethical issues include plagiarism, a risk of amplifying biases and inaccuracies, research fraud, copyright issues, and low transparency in content generation (Liebrenz et al., 2023; Sallam, 2023; Salvagno et al., 2023). These challenges may cause AICs to spread misinformation, with harmful consequences (Liebrenz et al., 2023; Sallam, 2023).

To better understand medical researchers’ attitudes and perceptions towards the use of AICs in the scientific process, we conducted a large-scale, international, cross-sectional survey. This survey sought to investigate the extent to which researchers are familiar with AICs, the perceived potential benefits and limitations of using these AICs in scientific research, and the factors that may influence their adoption.

## Methods

### Open Science Statement

This study’s protocol was registered on the Open Science Framework (OSF) (https://doi.org/10.17605/OSF.IO/SRM34) before respondent recruiting began. A protocol of this study was also preprinted on MedRxiv (https://doi.org/10.1101/2023.07.26.23293211). All study materials and cleaned data can be found on OSF (https://doi.org/10.17605/OSF.IO/25Y8Q).

### Ethical Considerations

Prior to conducting this study, ethics approval was obtained from the Ottawa Hospital Research Institute Research Ethics Board (REB). Informed consent was also obtained from all respondents prior to their participation in the survey. All data collected was kept confidential and anonymous, and no identifying information of respondents was collected.

### Study Design

We conducted an anonymous, cross-sectional, closed survey of corresponding authors of articles published in journals indexed in MEDLINE to investigate their attitudes and perceptions towards the use of AICs in the scientific process.

### Sampling Framework

A complete list of all journals indexed in MEDLINE (approximately 5300 journals as of April 2023) was obtained along with their NLM IDs. A search strategy, developed by JYN and reviewed by the rest of the team, of these NLM IDs was in OVID MEDLINE (**Appendix 1**, https://osf.io/7e8yz). The search was limited to records indexed over the two months prior to searching (2023/03/01-2023/04/07). This period was chosen as corresponding authors who have published between these dates were likely to still be actively involved in medical research and available to respond to a survey request. Authors of all article types were included. PubMed Identifier (PMID) numbers associated with all yielded articles were exported from OVID as a .csv file and inputted into an R script (created based on the easyPubMed package (easyPubMed, 2023)) to capture author names, affiliated institutions, and email addresses. A total of 122 323 PMIDs were retrieved and exported into R by SM and JYN. Corresponding author email extraction was completed on May 29, 2023. All generated results were combined into a master list and cleaned for potential errors/duplicates before survey administration.

### Inclusion Criteria

Respondents had to have met the following inclusion criteria: 1) self-identified as medical researchers (of any kind, whereby their research in one way or another contributes to the field of medicine), and 2) have completed at least one terminal degree in their respective field of study (e.g., PhD or equivalent in their respective field, MD or equivalent in their respective profession) or have >5 years of experience in a research-focused role (e.g., research coordinator). Students (both undergraduate and graduate) were ineligible to participate in this survey.

### Respondent Recruitment

As PMIDs were collected from all MEDLINE-indexed journals, a range of academic disciplines within the field of medicine were included (e.g., life sciences, public health, bioengineering, medical education). We used convenience sampling to recruit respondents, targeting corresponding authors identified by our sampling strategy. An email containing a recruitment script approved by the REB, detailing the study and its purpose, and a link to the survey was sent through email using the Microsoft Outlook Mail Merge software (Microsoft Outlook, 2023) on July 9, 2023. When respondents clicked on the survey link, they were directed to a webpage containing an informed consent form. Respondents had to agree to the form to proceed to the survey. This survey was closed, meaning only invited respondents were able to participate.

If respondents did not respond to the original invitation email, reminder emails were sent twice in batches of 10 000 during the weeks of July 17, 2023 and July 24, 2023. After deduplicating the list, 61 560 corresponding authors were sent invitations to participate. A 1-week waiting period followed the final email reminder to accommodate remaining interested respondents. Overall, respondents had 3 weeks to complete the survey which was closed on August 11, 2023.

Financial compensation was not offered; participation was entirely voluntary and anonymous. With the exception of the screening question, all other questions could be skipped if participants did not wish to respond. Participation could be withdrawn at any time during the survey simply by closing the browser, however, withdrawal was not possible after the survey was submitted given that the responses were collected anonymously.

### Survey

The complete survey can be found in **Appendix 2 (**https://osf.io/7gf6e). The survey was created, distributed, and collected using the University of Ottawa’s version of the SurveyMonkey software (University of Ottawa, 2023). The first draft of the survey was created by JYN, then reviewed by the rest of the team; the survey was developed based on a review of the literature and input from experts in the field of AI and scientific research. Survey questions were beta-tested by a team of authors and 2 invited researchers outside of the author list prior to administration of the survey. The survey contained 29 questions in total and took approximately 15 minutes to complete.

The survey consisted of both closed-ended (e.g., multiple choice, yes/no) and open-ended questions (i.e., respondents typed their answer in the text box provided), covering the following topics:

- Demographic information: age, gender, country of employment, level of education, publication record, and years of experience in medical research.
- Experience with AICs: extent to which respondents are familiar with AICs, their personal experience with AICs, and their likelihood of utilizing chatbots in the future.
- Role of AICs in the scientific process: respondents’ perceptions of AIC utilization in different steps of the scientific process, and the potential impact of chatbots on the future of scientific research.
- Perceived benefits of AICs in the scientific process: respondents’ perceptions of the potential benefits of using AICs in scientific research, such as increased efficiency, reduced human workload, and increased accuracy and dissemination.
- Perceived challenges of AICs in the scientific process: respondents’ perceptions of the potential challenges of using AICs in scientific research, such as ethical concerns, risk of bias, and potential inaccuracies.
- Open-ended questions: respondents had the opportunity to provide additional comments and feedback on the use of AICs in scientific research.

### Data Management and Analysis

Survey data was exported and analysed using Microsoft Excel. Counts and percentages were generated and used to summarize the collected data. Respondent demographic information was described. Qualitative data collected from open-ended questions was analysed thematically. Prior to beginning thematic analysis, pilot coding was conducted by two authors (AL and NS). Each author independently coded the responses of the first 20 respondents for question 14, and then collaborated to develop a shared codebook based on their results. After reaching a consensus on the code(s), all remaining open-ended responses were coded and the codebook was iteratively updated. Following this, individual codes were grouped into the themes by the two authors independently and finalized through consensus. Any conflicts were resolved through discussion between the two authors and if needed, by a third author (SGM). All data was then reviewed by JYN, followed by all remaining authors. A descriptive definition of each theme was then created.

## Results

Our collected raw survey data from SurveyMonkey can be found in **Appendix 3 (**https://osf.io/f62ds). Any personal identifiers provided by participants have been redacted.

### Respondent Demographics

Survey invitations were sent to a total of 61 560 email addresses, of which 2452 invitees provided responses (4.0% response rate). Of the respondents, 2292 met the eligibility criteria (93.5%), of whom 2165 completed the survey (94.5% completion rate).

As not all respondents completed all survey questions, the total number of responses for each question varies and is provided in parentheses throughout the presentation of results that follow.

Demographic data can be found in **Table 1**. For this question, the majority identified as male (n=1161 of 2149 respondents, 54.0%), less than half identified as female (n=959, 44.6%) and 29 respondents either identified as nonbinary, self-described, or opted not to say (1.3%). Most respondents fell within the 36 to 45 years age group (n=706/2156, 32.8%). Respondents self-identified as coming from 95 countries, with the greatest representation from the United States of America (n=601/2137, 28.1%), Canada (n=163, 7.6%), and Italy (n=129, 6.0%). Most were senior researchers (n=1136/2165, 52.5%), faculty members at a university/academic institution (n=1369/2158, 63.4%), primarily conducted clinical research (n=1107/2159, 51.3%), and had published more than 21 research articles (n=1549/2133, 72.4%).

**Table 1:** Respondent Demographic Data.

| Question | Answer Choices | Frequency | Percent |
| --- | --- | --- | --- |
| **Career stage** | Early career researcher (<5 years of formally starting your career post formal education) | 466 | 21.52% |
|  | Mid-career researcher (5-10 years of starting your career post formal education) | 563 | 26.00% |
|  | Senior researcher (>10 years of starting your career post formal education) | 1136 | 52.47% |
| **Total** |  | **2165** | **99.99%** |
| **Age** | 18-24 | 11 | 0.51% |
|  | 25-35 | 425 | 19.71% |
|  | 36-45 | 706 | 32.75% |
|  | 46-55 | 536 | 24.86% |
|  | 56-65 | 310 | 14.38% |
|  | 65+ | 153 | 7.10% |
|  | Prefer not to say | 15 | 0.70% |
| **Total** |  | **2156** | **100.00%** |
| **Gender** | Male | 1161 | 54.03% |
|  | Female | 959 | 44.63% |
|  | Non-binary | 10 | 0.47% |
|  | Prefer not to say | 16 | 0.74% |
|  | Prefer to self-describe, please specify. | 3 | 0.14% |
| **Total** |  | **2149** | **100.00%** |
| **Country of primary employment**<br>**(10 most popular shown)** | United States of America | 601 | 28.12% |
|  | Canada | 163 | 7.63% |
|  | Italy | 129 | 6.04% |
|  | United Kingdom of Great Britain and Northern Ireland | 113 | 5.29% |
|  | Australia | 111 | 5.19% |
|  | China | 86 | 4.02% |
|  | Spain | 74 | 3.46% |
|  | India | 61 | 2.85% |
|  | Netherlands | 61 | 2.85% |
|  | Brazil | 45 | 2.11% |
| **Total** |  | **1444** | **67.57%** |
| **Current position** | Faculty member at a university/academic institution | 1369 | 63.44% |
|  | Academic research staff (e.g., research coordinator) at a university/academic institution) | 370 | 17.15% |
|  | Government researcher | 95 | 4.40% |
|  | Clinician researcher | 470 | 21.78% |
|  | Pharmaceutical industry/company employee | 24 | 1.11% |
|  | Scholarly communications employee (e.g., scholarly journals/publishing, medical writing) | 10 | 0.46% |
|  | Consultant (e.g., own or employed by research consulting firm) | 64 | 2.97% |
|  | Third sector (E.g., NGO, non-profit) | 53 | 2.46% |
|  | Other (please specify) | 153 | 7.09% |
| **Total** |  | **2158** | **100.00%** |
| **Primary research area** | Clinical research | 1107 | 51.27% |
|  | Preclinical research – in vivo | 349 | 16.16% |
|  | Preclinical research – in vitro | 355 | 16.44% |
|  | Health systems research | 260 | 12.04% |
|  | Health services research | 392 | 18.16% |
|  | Methods research | 333 | 15.42% |
|  | Epidemiological research | 549 | 25.43% |
|  | Other (please specify) | 295 | 13.66% |
| **Total** |  | **2159** | **100.00%** |
| **Number of research articles published to date** | ≤2 | 30 | 1.41% |
|  | 3-10 | 275 | 12.89% |
|  | 11-20 | 279 | 13.08% |
|  | 21-50 | 548 | 25.69% |
|  | 51-100 | 424 | 19.88% |
|  | 101-200 | 359 | 16.83% |
|  | 201-500 | 189 | 8.86% |
|  | 501-800 | 19 | 0.89% |
|  | 801-1000 | 6 | 0.28% |
|  | 1001-1300 | 2 | 0.09% |
|  | Other (incomplete) | 2 | 0.09% |
| **Total** |  | **2133** | **100.00%** |

### Experience with AI Chatbots

Full details about respondent’s experience with AICs and their affiliated institution’s views on using AICs in the scientific process can be found in **Table 2**. Notable findings include that 1294/2138 respondents (60.5%) were familiar with the concept of AICs, and of those who had used AICs, most used ChatGPT (n=1402/2136, 65.6%). Over half of respondents used an AIC for purposes relating to the scientific process (n=1107/2125, 52.1%), indicated ‘yes’ and 882/1107 (80.0%) utilized ChatGPT for these purposes. However, 1180 respondents had never used an AIC in this context (55.5%). Respondents were likely (n=790/2136, 37.0%) or very likely to use AICs for their future research (n=451, 21.1%).

**Table 2:** Respondent Experience with AI Chatbots.

| Question | Answer Choices | Frequency | Percent |
| --- | --- | --- | --- |
| **How familiar are you with the concept of artificial intelligence chatbots?** | Very familiar | 283 | 13.24% |
|  | Familiar | 1294 | 60.52% |
|  | Unfamiliar | 357 | 16.70% |
|  | Very unfamiliar | 169 | 7.90% |
|  | I'm not sure | 35 | 1.64% |
| **Total** |  | **2138** | **100.00%** |
| **Which of the following AI chatbots have you used before (for any purpose)? (Please select all that apply).** | Bing Chat | 246 | 11.52% |
|  | ChatGPT | 1402 | 65.64% |
|  | YouChat | 26 | 1.22% |
|  | Google Bard | 206 | 9.64% |
|  | I have never used an AI chatbot | 658 | 30.81% |
|  | Other (please specify) | 75 | 3.51% |
| **Total** |  | **2136** | **100.00%** |
| **Which of the following AI chatbots have you used before specifically for purposes relating to scientific processes? (Please select all that apply).** | Bing Chat | 87 | 4.09% |
|  | ChatGPT | 882 | 41.51% |
|  | YouChat | 9 | 0.42% |
|  | Google Bard | 82 | 3.86% |
|  | I have never used an AI chatbot for purposes relating to scientific processes | 1180 | 55.53% |
|  | Other (please specify) | 47 | 2.21% |
| **Total** |  | **2125** | **100.00%** |
| **How likely are you to use an AI chatbot for your research in the future?** | Very likely | 451 | 21.11% |
|  | Likely | 790 | 36.99% |
|  | Unlikely | 348 | 16.29% |
|  | Very unlikely | 248 | 11.61% |
|  | I am not sure | 299 | 14.00% |
| **Total** |  | **2136** | **100.00%** |
| **Does your research institution provide any training on how to appropriately use AI tools in the scientific process?** | Yes, and I have taken it | 64 | 2.99% |
|  | Yes, but I have not taken it | 180 | 8.42% |
|  | No | 1487 | 69.58% |
|  | I am not sure | 406 | 19.00% |
| **Total** |  | **2137** | **100.00%** |
| **Has your research institution implemented any policies surrounding the use of AI chatbots in the scientific process?** | Yes | 211 | 9.90% |
|  | No | 1178 | 55.28% |
|  | I am not sure | 742 | 34.82% |
| **Total** |  | **2131** | **100.00%** |

Most respondents reported that their research institution did not offer any training on using AI tools (n=1487/2137, 69.6%), though 244/2137 (11.4%) indicated that their institution offered training on appropriate use of AI tools in the scientific process while only 64/244 (26.2%) completed such training. When asked if their institution had implemented any policies surrounding AIC use in the scientific process, 211/2131 respondents (10.0%) answered yes.

### Role of AI Chatbots in the Scientific Process

Full participant responses about their perceived role(s) of AICs in the scientific process can be found in **Table 3**. Over half of respondents answered that some training is necessary (n=1048/2049, 51.2%), while 665 (32.5%) felt a lot of training was necessary to effectively use AICs in the scientific process. Most respondents expressed clear interest in learning more/receiving training on using AICs in the scientific process (n=1428/2048, 69.7%). Most respondents felt AICs would be very important (n=741/2050, 36.2%) or important (n=935, 45.6%) in the future of scientific research. Most respondents felt AICs would have a very positive or positive outcome (n=1236/2048, 60.4%) while 380 (18.6%) felt AICs would negatively or very negatively impact future scientific research.

**Table 3:** Role of AI Chatbots in the Scientific Process.

| **Question** | **Answer Choices** | **Frequency** | **Percent** |
| --- | --- | --- | --- |
| **How much training and education do you think researchers need in order to effectively use AI chatbots in the scientific process?** | A lot | 665 | 32.45% |
|  | Some | 1048 | 51.15% |
|  | Very little | 173 | 8.44% |
|  | None | 34 | 1.66% |
|  | I am not sure | 129 | 6.30% |
|  | **Total** | **2049** | **100.00%** |
| **Would you be interested in learning more/receiving training about how to use AI chatbots in the scientific process?** | Yes | 1428 | 69.73% |
|  | No | 189 | 9.23% |
|  | Maybe | 431 | 21.04% |
|  | **Total** | **2048** | **100.00%** |
| **How important do you think AI chatbots will be in the future of scientific research?** | Very important | 741 | 36.15% |
|  | Important | 935 | 45.61% |
|  | Unimportant | 110 | 5.37% |
|  | Very unimportant | 18 | 0.88% |
|  | I'm not sure | 246 | 12.00% |
|  | **Total** | **2050** | **100.00%** |
| **In general, how do you perceive the potential impact of AI chatbots on the future of scientific research?** | Very positively | 282 | 13.77% |
|  | Positively | 954 | 46.58% |
|  | Negatively | 290 | 14.16% |
|  | Very negatively | 90 | 4.39% |
|  | I am not sure | 432 | 21.09% |
|  | **Total** | **2048** | **100.00%** |

Respondents rated their agreement with statements regarding how helpful an AIC would be in different steps of the scientific process, on a 5-point scale ranging from “Very helpful” to “Very unhelpful”. A graph detailing full responses is depicted in **Figure 1**. Notably, 1235/2044 respondents (60.4%) felt AICs would be very helpful or helpful for conducting literature searches. Respondents had mixed views regarding AIC helpfulness in understanding/selecting a research methodology; 820/2033 (40.3%) perceived AICs to be unhelpful or very unhelpful, whereas 754/2033 (37.1%) felt they would be very helpful or helpful. Of the participants who answered this question, senior researchers tended to use it *always, often, sometimes* compared to other researchers in different career stages. The findings for the results stratified by participants’ usage of AI tools in scientific processes can be found in **Appendix 4 (**https://osf.io/v2csn). Respondents overwhelmingly felt that AICs would be very helpful or helpful when writing/editing manuscripts (n=1329/2040, 65.2%), research grant applications (n=1183/2032, 58.2%), and when translating research materials into another language (n=1346/2044, 65.9%). Of 2039 respondents, 902 (44.2%) perceived AICs to be either unhelpful or very unhelpful for peer review/critiques. Eight hundred and forty-eight of 2041 respondents (41.6%) felt AICs would be very helpful or helpful for generating presentation materials, while 1059/2040 respondents(51.9%) thought AICs would be very helpful or helpful for research engagement. Lastly, 1261/2040 respondents (61.8%) believed AICs were very helpful or helpful for performing general administrative tasks.

**Figure 1:**
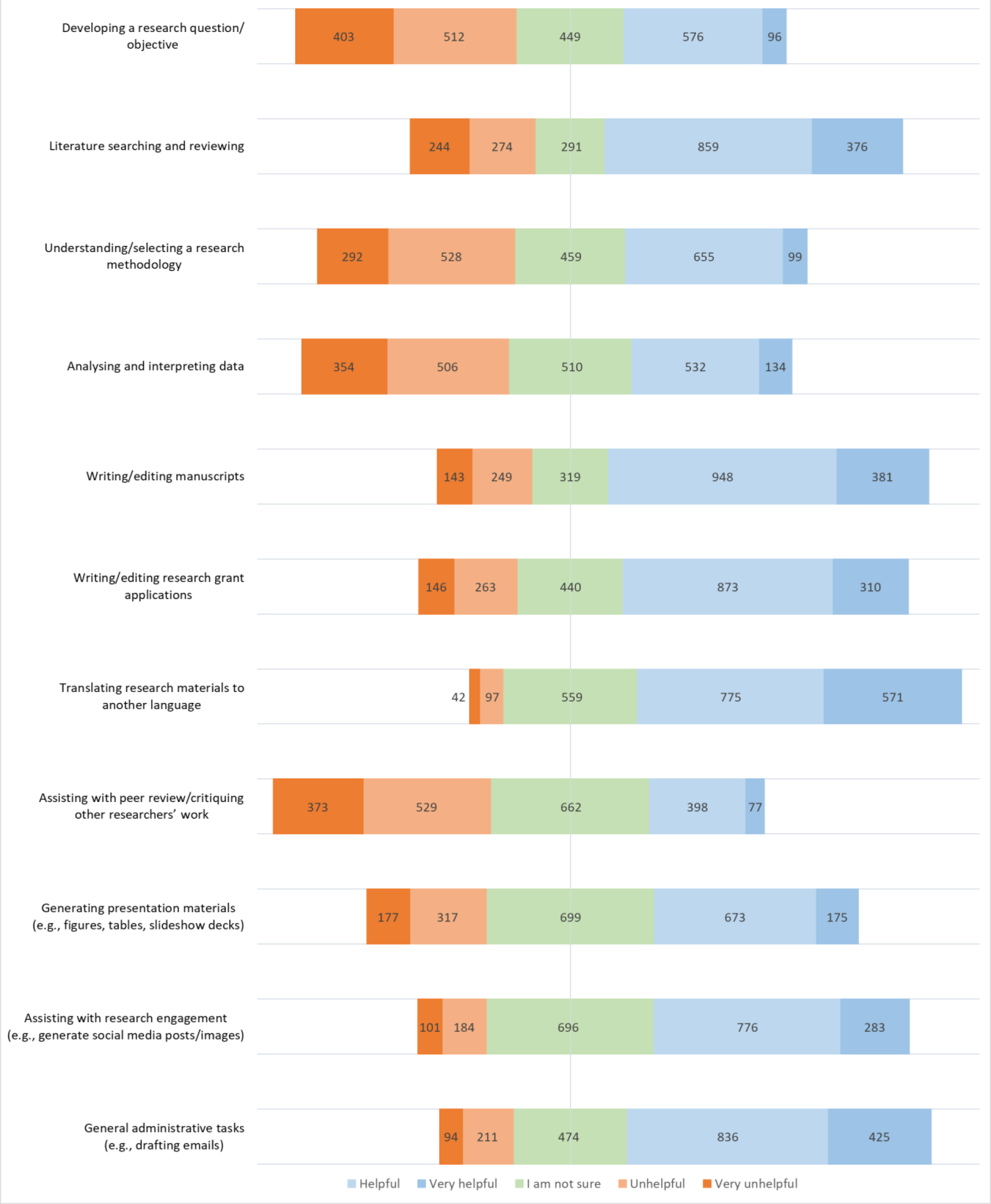
Respondent Agreement with Statements Regarding the Helpfulness of AI Chatbots in Different Steps of the Scientific Process.

Respondents rated how often they had used AICs in different steps of the scientific process, on a 5-point scale ranging from “Always” to “Never”. Frequencies and percentages of these responses are depicted in **Figure 2**. Notable findings include that 477/893 (53.4%) had sometimes or rarely used an AIC for literature searches and reviews, 370/896 (41.3%) sometimes or rarely used an AIC to write/edit manuscripts, and 315/895 respondents (35.2%) sometimes or rarely used an AIC for writing/editing research grant applications. Regarding language translation, 461/895 (51.5%) had never used AICs for this purpose; however of those who did use an AIC for translation purposes (n=434/895, 48.5%), the top five countries found AI to be very helpful: 1) United States of America (19.09%), 2) Italy (8.41%), 3) China (7.36%), 4) Canada (6.13%) and 5) Spain (1.08%). The findings for the results stratified by participants’ perceptions of AI helpfulness as a translational tool can be found in **Appendix 5 (**https://osf.io/kc4zt).

**Figure 2:**
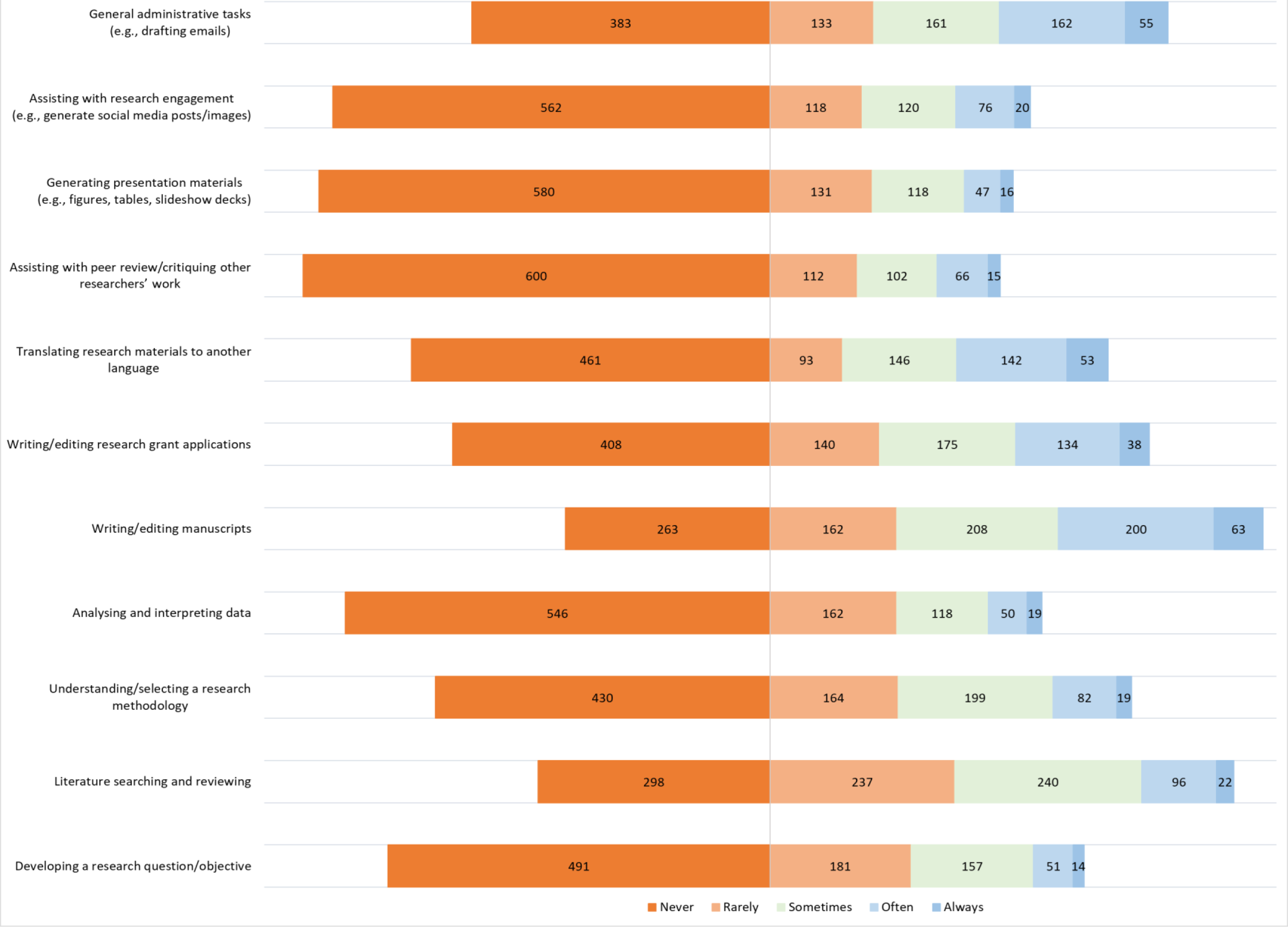
Respondent Use of AI Chatbots in Different Steps of the Scientific Process.

### Perceived Benefits and Challenges of AI Chatbots in the Scientific Process

Respondents rated their agreement with statements relating to how beneficial AICs could be in different steps of the scientific process, on a 5-point scale ranging from “Strongly disagree” to “Strongly agree”. Full details of these responses are depicted in **Figure 3**. The most agreed upon benefits of AICs included increased efficiency and speed of data analysis (n=1026/1939, 52.9%), reduction of workloads and administrative burdens on researchers (n=1299/1941, 66.9%), enabling of constant accessibility to scientific information and assistance (n=1081/1939, 55.7%), more effective handling and analysis of large datasets than human researchers (n=1086/1935, 56.1%), a cost-effective solution to conducting scientific research (n=974/1933, 50.4%), a way to provide a more inclusive research environment (n=1120/1936, 57.9%), and improved quality and efficiency of scientific communication and dissemination (n=1002/1932, 51.9%).

**Figure 3:**
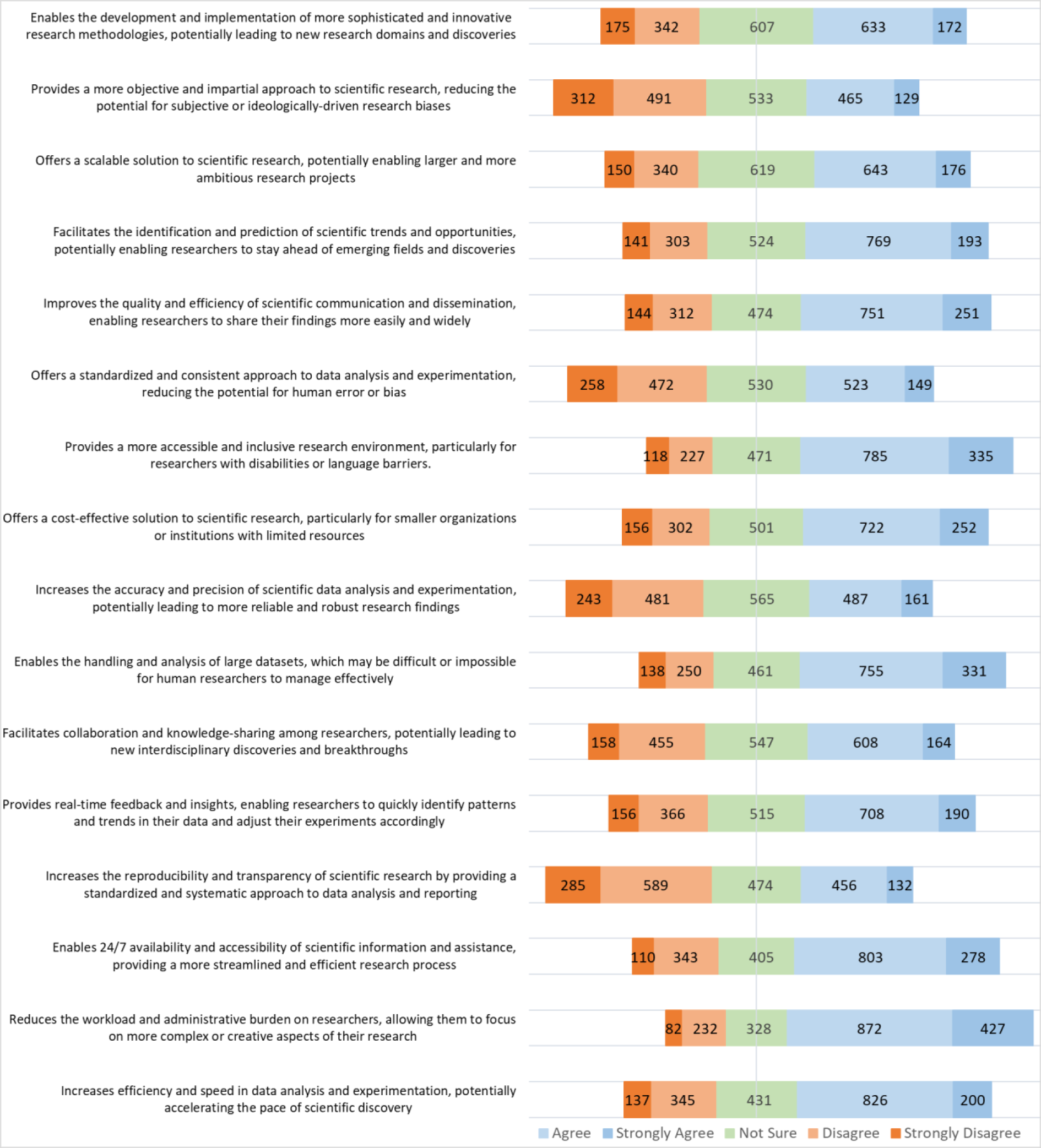
Respondent Agreement with Potential Benefits of Using AI Chatbots in the Scientific process.

Ratings on whether AICs could increase the reproducibility and transparency of research were mixed; 285/1936 participants strongly disagreed (14.72%) or disagreed (n=589, 30.4%), 474 (24.5%) were unsure, and 456 (23.6%) agreed or strongly agreed (n=132, 6.8%) with the statement. Five hundred and sixty-five participants of 1937 (29.2%) were unsure if AICs could increase the accuracy and precision of data analysis and experimentation, whereas 243 respondents (12.6%) strongly disagreed or disagreed (n=481, 24.8%) with the statement.

Respondent views were mixed on whether AICs could reduce human error or bias by providing a standardized approach to data analysis; 530 of 1932 (27.4%) were unsure, 523 (27.1%) agreed, and 149 (7.7%) strongly agreed, however 472 respondents (24.4%) disagreed, and 258 respondents (13.4%) strongly disagreed with the statement. Respondents rated their agreement with statements relating to potential challenges of using AICs in the scientific process, on a 5-point scale ranging from “strongly disagree” to “strongly agree”. Full responses are depicted in **Figure 4**. The most agreed upon challenges of AICs included biased/skewed data outputs (n=1384/1924, 71.9%), ethical and legal concerns (n=1480/1927, 76.8%), resistance/pushback from researchers (n=1374/1924, 71.4%), lack of accountability (n=1405/1924, 73.0%), lack of transparency and interpretability in their decision-making process (n=1456/1923, 75.7%), lack of understanding in how they make decisions and generate responses (n=1484/1923, 77.2%), limited in handling context-dependent or situation-specific information (n=1426/1924, 74.1%), limited in capturing the nuances and complexities of human thought and reasoning (n=1467/1921, 76.4%), data privacy and security concerns (n=1401/1921, 72.9%), and challenges related to user acceptance and adoption (n=1437/1924, 74.7%).

**Figure 4:**
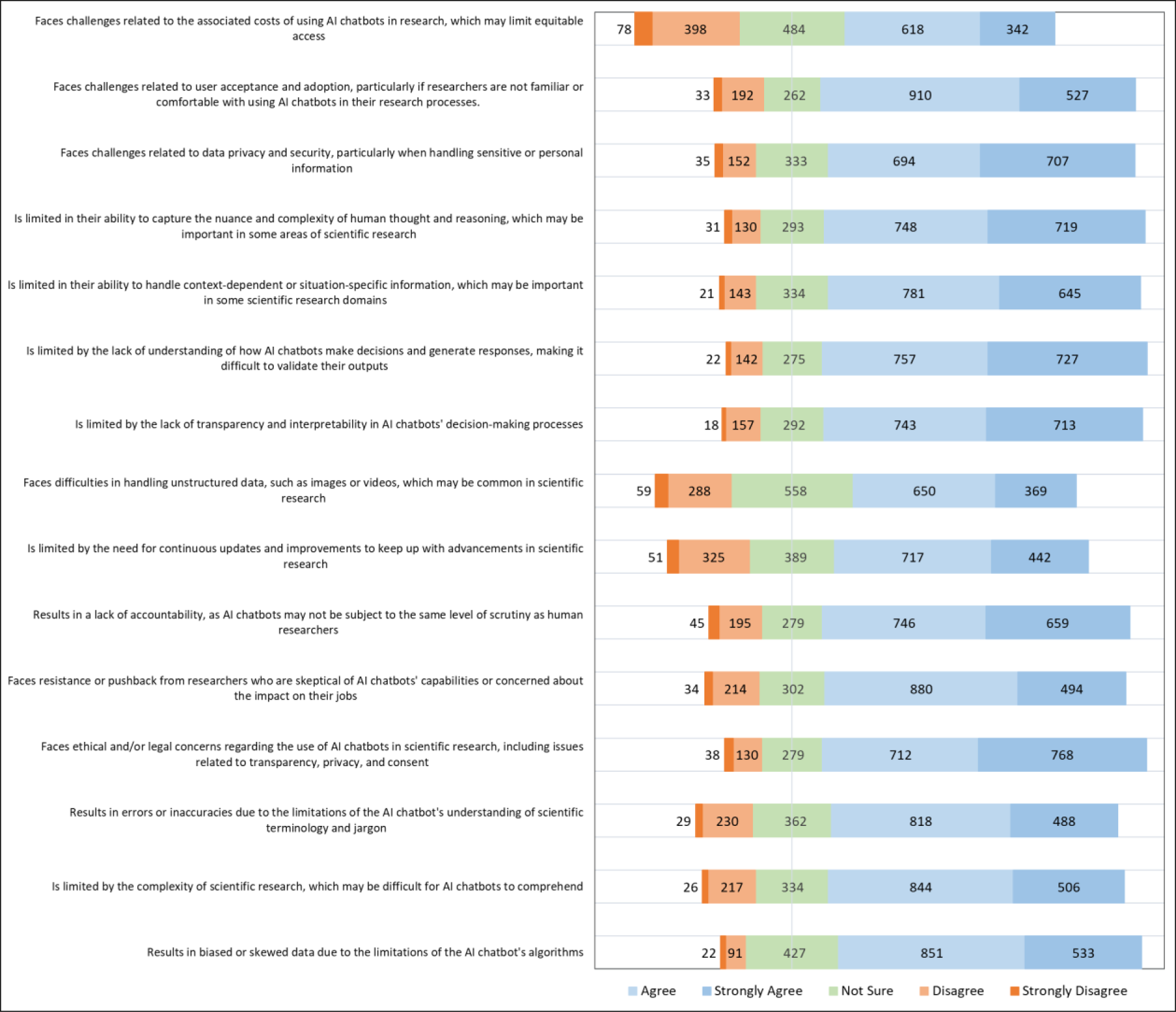
Respondent Agreement with Potential Challenges of Using AI Chatbots in the Scientific Process.

### Open-ended Questions

The open-ended questions allowed participants to provide additional feedback and comments on the use of AICs in scientific research. The open-ended questions’ designated themes, sub-themes, individual codes, and their frequencies can be found in **Appendix 6 (**https://osf.io/7qfc3). Full results to the thematic analysis tallies can also be found in **Appendix 7 (**https://osf.io/k67cp).

### Nature of AI Chatbot Training and Its Composition

From Question 14 regarding the nature of AIC training and its components, three major themes emerged: 1) guidance, 2) benefits/disadvantages of using AICs in research, and 3) legal affairs. The most prevalent theme was ‘guidance’, which entailed responses that described guidelines received from institutions on AICs in scientific research. The most common sub-themes under ‘guidance’ included ‘how AI chatbots work’ and ‘structured/formal training provided’.

### Nature of AI Chatbot Policies in Research Institutions

From Question 16 regarding the nature of AIC policies in research institutions, four major themes emerged: 1) guidance, 2) ethics, 3) legal affairs, and 4) benefits/disadvantages of using AICs in research. The most prevalent themes were ‘guidance’ and ‘ethics’ which described both general and ethical guidelines received from institutions on AICs respectively. The most common sub-themes were ‘plagiarism in AI’ under the theme ‘ethics’ and ‘security and confidentiality’ under the theme ‘legal affairs’.

### Helpfulness of AI Chatbots in Additional Steps of the Scientific Process

From Question 18 regarding the helpfulness of AICs in additional steps of the scientific process, two prevalent themes emerged: 1) writing aids and 2) other. The theme ‘writing aids’ described responses that mention using AICs for writing anything within the scientific process. In contrast, the theme ‘other’ described responses that expressed the respondent’s uncertainty or lack of knowledge regarding the answer to the question.

### Extent of AI Chatbot Usage in Additional Steps of the Scientific Process

From Question 20 regarding the extent of AIC usage in additional steps of the scientific process, two prevalent themes emerged: 1) writing aids and 2) programming. The theme ‘writing aids’ described responses that mention using AICs for writing, reviewing, editing, or correcting the English language for enhanced clarity. The theme ‘programming’ described responses that expressed the ability of AICs to optimize programming tasks.

### Additional Benefits of AI Chatbots in the Scientific Process

From Question 26 regarding the additional benefits of AICs in the scientific process, two prevalent themes emerged: 1) writing aids and 2) other. The theme ‘writing aids’ described responses that mention using AICs for writing, reviewing, editing, correcting the English language for enhanced clarity, and discussing figure generations. In contrast, the theme ‘other’ described responses that expressed the respondent’s uncertainty in providing an answer to the question.

### Additional Challenges to the Use of AI Chatbots in the Scientific Process

From Question 28 regarding the additional challenges to the use of AICs in the scientific process, two prevalent themes emerged: 1) reliability and 2) ethical issues. The theme ‘reliability’ described responses that question the trustworthiness of AIC generated information. The theme ‘ethical issues’ described responses that questioned the ethical aspects of using AI in research.

### Final Comments Regarding AI Chatbot Use or the Survey

From Question 29 regarding the additional challenges to the use of AICs in the scientific process, two prevalent themes emerged: 1) other and 2) limitations in AI. The theme ‘other’ included responses that were not relevant to the purpose of the research (e.g., feedback on the survey). The theme ‘limitations in AI’ described responses that expressed doubt in the design and implementation of AI.

## Discussion

### Significance of Findings

To our knowledge, this survey is the first of its kind and of this magnitude to provide an international perspective on medical researcher knowledge, use, and perceptions of AICs in the scientific process and their potential impact on how medical research is conducted at a time when these AICs are newly being applied to the field. Overall, respondents expressed mixed opinions regarding the potential benefits of using AICs in the scientific process, whereas most respondents agreed upon the disadvantages and challenges of utilizing these AICs. Many agreed that AICs could be helpful in various steps of the scientific process, but not in their current state due to their many limitations and errors.

In our survey, most respondents were already familiar with the concept of an AIC and had utilized an AIC before for research purposes, with the most utilized AIC being ChatGPT. Respondents overwhelmingly felt AICs were most helpful in writing and editing manuscripts. However, only 244 respondents reported that their institution offered training on the appropriate use(s) of AI tools/chatbots in the scientific process and 211 reported that their affiliated institution implemented policies regarding the use of AI tools/chatbots in scientific research. Considering that many widely available AICs still have major and well-known shortcomings (e.g., hallucinations), it is both surprising and unfortunate that awareness campaigns addressing the limitations of AI were only offered in a handful of research institutions and that so few institutions had implemented clear policies on AICs use in research. The lack of available training and policies could be due to the relative newness of AIC use in research or perhaps due to the prohibition of using AI tools, like chatbots, in research articles by several publishing organizations and journals (Brainard, 2023; Lee, 2023). It certainly is not for lack of interest, as an overwhelming majority of the respondents conveyed clear interest in learning more and receiving training about using AICs in the scientific process.

This lack of training and policies also raises concerns for research integrity, as many journals now require authors to disclose all AI use, yet it is known that undeclared ChatGPT-assisted manuscripts are being published in peer-reviewed journals (Conroy, 2023). Not only does this indicate a lack of effective safeguarding tools/mechanisms surrounding the detection of AI-generated material in the peer review process, but also a worrying pattern that research integrity may already be compromised.

The principal perceived benefit of AICs is time saved in drafting manuscripts, language translation, or generating figures and tables. AICs are already known for their ability to automate time-consuming tasks in the research process and their ability to provide writing and editing assistance (Sallam, 2023; Salvagno et al., 2023); benefits that are also reflected by our findings.

Interestingly, despite respondents viewing AICs as generally helpful in various steps of the scientific process, few have actually used them. Most respondents reported having never used an AIC due to significant limitations still present in many current AIC models. Respondents were most concerned about the difficulty in validating AIC outputs due to the lack of understanding/transparency behind their decision-making and response generation processes.

### Comparative Literature

The use of AI tools has already had much discussion in medicine and healthcare due to their pattern recognition capabilities when handling medical data. A systematic review by Ali et al. (2023) found that the benefits to patients of AI use in healthcare include early diagnosis, patient monitoring, and automated decision-making. Goodman et al. (2023) state that AI tools incorporated into clinical decision support (CDS) systems have guided clinical decision making to make algorithmic predictions under conditions of clinical uncertainty using patient health records. For example, sepsis warning systems can use real-time data to identify early-stage sepsis in patients before physicians can even detect any clinical signs (Goodman et al., 2023). Physicians can therefore incorporate AI-powered CDS predictions into their clinical decisions; however, these predictions require critical evaluation and human judgement as the AI algorithm contains bias and can also produce false positives (Goodman et al., 2023). Similarly, Ruksakulpiwat et al. (2023) state that although ChatGPT can provide general and basic-level medical information and simplify medical reports for radiologists quite well, it is not without errors and currently requires human validation. In our survey, many respondents also noted that AICs can provide real-time feedback and insights, be useful in data analysis— particularly in large datasets—and facilitate the identification and prediction of scientific trends and opportunities. Survey respondents also emphasized that if AICs were to be used in scientific research, the outputs of these AICs require human judgement and evaluation due to the many biases and errors they contain.

Many AICs are now easily accessible and available at little to no cost for an initial use to the public. A 2023 poll conducted by YouGov found that 62% of American respondents were somewhat or mostly concerned about the growth of artificial intelligence (Heath, 2023). As well, 56% of respondents expressed support for federal regulation of AI over self-regulation by tech companies, as 82% of respondents did not trust tech executives to regulate AI (Heath, 2023). These findings are also supported by Gillespie et al. (2023), who conducted a global survey of over 17 000 people and found that 61% of respondents were wary about trusting AI systems. Respondents were most concerned about the safety, security, data privacy measures, and fairness of AI systems (Gillespie et al., 2023), which were among the principal concerns voiced by our survey respondents for using AICs in scientific research.

Regarding AIC use in research, a 2023 survey conducted by User Interviews aimed at UX researchers/designers and Research Ops specialists found that 77.1% of respondents were already using AI in at least some of their work (Burnam, 2023). About half (51.1%) used ChatGPT in their research. Burnam (2023) reports that the most-cited benefit of AI was its efficiency, as 40.4% of respondents used AI for qualitative coding purposes and 45.5% used AI for writing reports. A survey of 1600 researchers by Nature (2023) also supported these perceived benefits, as respondents felt AI tools in research would increase the processing of data, speed up computations, automate data acquisition, and reduce overall time and resources spent by researchers. In addition, negative effects of AI use in research were perceived to be primarily the risk of bias/discrimination in data (55%), fraud, (55%) and increased reliance on pattern recognition without understanding (69%) (Noorden & Perkel, 2023).

### Implications

In our survey, many researchers indicated they had already used an AIC for research purposes, but most were also never formally trained as their affiliated institutions did not offer such training or have policies regarding their use in place. Most researchers also wanted to learn more about and receive training on using AICs in scientific research. This suggests a need to develop structured training for biomedical researchers on using AICs in the scientific process as many want to and are already using widely available AICs without formal training. Creating set guidelines and standardized training surrounding AIC use in research may help to maintain research integrity. Many researchers also expressed concern over AIC use in research due to the many limitations/errors and ethical issues current models perpetuate. Before these tools can be used in research, these limitations and concerns need to be addressed. Lastly, AICs can potentially enhance research equity by helping non-native speakers overcome language barriers (Liebrenz et al., 2023; Sallam, 2023; Salvagno et al., 2023), given that these AICs continue to be widely available for free or at a low-cost.

### Future Directions

This survey provides insight into early AIC use in medical research, which can be used to measure this technology’s evolution. Currently, there are significant concerns and challenges regarding AIC usage that need urgent attention. Future research should centre around developing guidelines and formal training surrounding appropriate AIC use in scientific research and to facilitate a greater understanding of these tools. Furthermore, a code of ethics concerning the use of AI tools, like AICs, in academia and research is needed (Sallam, 2023; Salvagno et al., 2023), considering the potential impact improper use may have on researchers, institutions, education, and the publishing industry, and the potential benefits appropriate use can offer towards the dissemination of knowledge. Furthermore, current limitations and errors in AIC models need to be addressed by AIC developers.

### Strengths and Limitations

As cross-sectional surveys only observe a population at one point in time, they are relatively quick and inexpensive to conduct. An additional strength of our study is that only names and email addresses of corresponding authors from the most recent 2 months were retrieved to minimize bounce back/inactive emails.

Our study has several limitations. non-English speaking researchers are largely excluded from our sample due to our own language and resource limitations. In addition, respondents who are only partially fluent in English may find some difficulty in participating in our survey. This would likely exclude their perceptions and attitudes; thus, our findings may not be applicable to those who primarily publish their research in languages other than English. Another limitation is that our raw response rate will be underestimated as we did not account for bounce back/inactive email accounts and autoreplies or determine how many of the 61 560 authors emailed identified as a medical researcher Additionally, although we aimed to assess the thoughts of a large sample of researchers from different disciplines, many invitees expressed that they could not participate as they had never used AICs previously. In such instances, one of us (JYN) encouraged these researchers to still take the survey. Despite our best efforts to encourage all invitees, regardless of AIC experience, to participate in the survey, the generalizability of our findings may be affected as those with either strong opinions for or against the use of AICs were more likely to respond to our survey. Thus, those with little experience or those who are unsure about the use of AICs may be underrepresented in our survey. While the relatively low response rate from this pool impacts generalizability, given the scale of our sampling, the final total number of participants still represents a relatively large sample size. Another limitation is that AIC use in research can only be observed at one point in time, therefore, our results will only provide a snapshot of researchers’ perspectives at an early stage of AIC use. Inherent to the cross-sectional survey study design, our study is also susceptible to recall and non-response bias.

## Data Availability

All data and materials associated with this study is available in this manuscript or has been made publicly available for download on the Open Science Framework.

https://doi.org/10.17605/OSF.IO/25Y8Q

## List of Abbreviations

AI: artificial intelligence
AIC: artificial intelligence chatbot
CDS: clinical decision support
ChatGPT: chat generative pre-trained transformer
COPE: Committee on Publication Ethics
LLM: large language model
OSF: Open Science Framework
PMID: PubMed Identifier
WAME: World Association of Medical Editors

## Declarations

### Ethics Approval and Consent to Participate

Ethics approval was granted by the Ottawa Health Science Network Research Ethics Board (REB Number: 20230288-01H).

### Consent for Publication

All authors consent to this manuscript’s publication.

### Availability of Data and Materials

All data and materials associated with this study is available in this manuscript or has been made publicly available for download on the Open Science Framework: https://doi.org/10.17605/OSF.IO/25Y8Q

### Authors’ Contributions

JYN: designed and conceptualized the study, co-drafted the manuscript, and gave final approval of the version to be published.

SGM: co-drafted the manuscript, and gave final approval of the version to be published.

AL: analysed data, made critical revisions to the manuscript, and gave final approval of the version to be published.

NS: analysed data, made critical revisions to the manuscript, and gave final approval of the version to be published.

CL: assisted with the design of the study and the analysis of data, made critical revisions to the manuscript, and gave final approval of the version to be published.

AI: assisted with the design of the study and the analysis of data, made critical revisions to the manuscript, and gave final approval of the version to be published.

RBH: assisted with the design of the study and the analysis of data, made critical revisions to the manuscript, and gave final approval of the version to be published.

DM: assisted with the design of the study and the analysis of data, made critical revisions to the manuscript, and gave final approval of the version to be published.

## Appendices Legend

**Appendix 1:** OVID MEDLINE Search Strategy Derived from All Journals Currently Indexed in MEDLINE, available at: https://osf.io/7e8yz

**Appendix 2:** Survey, available at: https://osf.io/7gf6e

**Appendix 3:** Raw Survey Data in the Artificial Intelligence Chatbots in the Scientific Process A Cross-Sectional Survey, available at: https://osf.io/f62ds

**Appendix 4:** Responses Categorized Based on Participants’ Usage of AI tools in Scientific Processes, available at: https://osf.io/v2csn

**Appendix 5:** Responses Categorized Based on Participants’ Perceptions of AI Helpfulness as a Translational Tool, available at: https://osf.io/kc4zt

**Appendix 6:** Raw Master List of the AI Thematic Analysis Coding, available at: https://osf.io/7qfc3

**Appendix 7:** Master List of Thematic Analysis Tally Sheets, available at: https://osf.io/k67cp

